# Epidemiology of Acute Sports-Related Digit Injuries in Young Athletes

**DOI:** 10.64898/2026.05.03.26352306

**Authors:** Shinsuke Sakoda

## Abstract

**Objectives:** To investigate the epidemiology of acute sports-related upper-extremity injuries in young athletes, with a particular focus on the frequency, anatomical distribution, injury types, and mechanisms of digit injuries.

**Methods:** This single-center retrospective observational study included athletes aged ≤22 years who sustained acute sports-related upper-extremity injuries between January 2017 and November 2025. Digit injuries were defined as injuries involving the thumb and fingers at or distal to the metacarpophalangeal joint. Injury characteristics, mechanisms, and sports categories were analyzed using descriptive statistics.

**Results:** A total of 1,219 acute sports-related upper-extremity injuries were analyzed. Digit injuries were the most common injury location, accounting for 412 cases (33.8%), followed by shoulder (30.7%), elbow (17.5%), wrist (14.4%), and palm injuries (3.6%). Jammed finger was the most frequent injury type, comprising 64.8% of digit injuries, followed by fractures (20.1%) and dislocations (5.3%). Most injuries were caused by contact mechanisms (90.3%), with ball contact being the predominant cause (49.5%). Ball sports accounted for 85.4% of all digit injuries.

**Conclusions:** Digit injuries represent the most frequent acute sports-related upper-extremity injuries in athletes aged ≤22 years, with jammed finger accounting for the majority of cases. Most injuries were associated with ball contact, highlighting the need for preventive strategies and appropriate initial management for digit injuries in young athletes.

## Introduction

Upper-extremity sports injuries are common among young athletes and can substantially affect not only sports participation but also physical development during the growth period (1,2). The upper extremity is frequently exposed to direct external forces in many sports, and injuries in this region may lead to decreased athletic performance and time loss from sports participation (1).

However, most epidemiological data on digit injuries have been derived from general trauma populations or emergency department–based studies, rather than from cohorts specifically focused on young athletes with acute sports-related injuries (3,4). Consequently, the actual burden and characteristics of digit injuries occurring in sports settings among young athletes remain insufficiently understood (4).

Among digit injuries, so-called jammed finger is often perceived as a relatively minor condition and therefore receives limited attention in clinical practice and research (5,6). Nevertheless, jammed finger represents a common injury pattern during sports activities, and clarification of its frequency, injury mechanisms, and anatomical characteristics is essential for appropriate initial management and the development of preventive strategies (5).

The purpose of this study was to investigate the epidemiology of acute sports-related upper-extremity injuries in athletes aged ≤22 years, with a particular focus on the frequency, anatomical distribution, injury types, and mechanisms of digit injuries.

## Methods

### Study Design

This study was a single-center retrospective observational study conducted at a regional tertiary care hospital.

### Participants

Athletes aged ≤22 years who visited the specialized outpatient clinic for sports-related injuries at our institution between January 2017 and November 2025 were eligible for inclusion. The study population consisted of patients who sustained acute upper-extremity injuries occurring during sports activities or directly attributable to sports participation.

Exclusion criteria were non–sports-related injuries, chronic or overuse conditions, duplicate visits for the same injury, incomplete medical records, and insufficient imaging data. Only one injury per athlete was included in the analysis.

A total of 1,219 acute sports-related upper-extremity injuries were ultimately included in the study.

### Definitions and Classification

Digit injuries were defined as injuries involving the thumb and fingers at or distal to the metacarpophalangeal (MP) joint. Injuries involving the palm and wrist were excluded from the digit injury analysis and classified separately.

Jammed finger was defined as a ligamentous or capsular injury without radiographic evidence of fracture on plain radiographs. Volar plate avulsion injuries with associated bony fragments were classified as fractures.

### Outcome Measures

The following variables were evaluated:

1. Anatomical distribution of upper-extremity injuries
2. Distribution of digit injuries by digit and joint level
3. Injury type
4. Injury mechanism
5. Sports category

### Statistical Analysis

Statistical analysis was performed using descriptive statistics. Categorical variables were summarized as counts and percentages (n, %).

### Ethical approval

This study was approved by the institutional review board of Ashiya Central Hospital, and informed consent was obtained using an opt-out method.

## Results

### Distribution of Upper-Extremity Injuries

A total of 1,219 acute sports-related upper-extremity injuries in athletes aged ≤22 years were analyzed. The mean age of the study population was 15.1 ± 3.7 years, and males accounted for 83.5% of the cases.

Digit injuries were the most common, comprising 412 cases (33.8%), followed by shoulder injuries (374 cases, 30.7%), elbow injuries (213 cases, 17.5%), wrist injuries (176 cases, 14.4%), and palm injuries (44 cases, 3.6%) (Table 1).

**Table 1.**
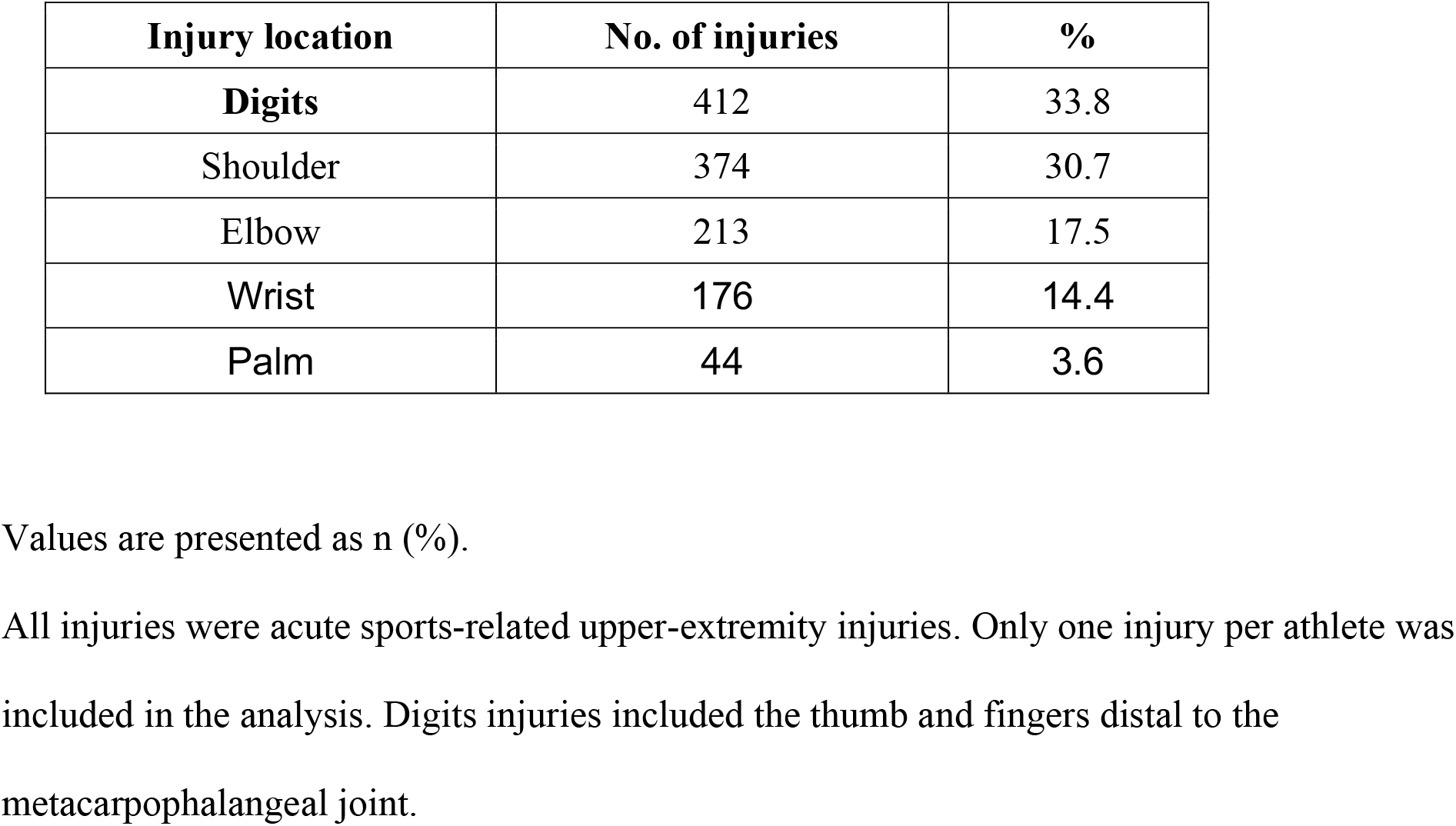
Distribution of acute sports-related upper-extremity injuries by anatomical location in athletes aged ≤22 years (n = 1219)

| Injury location | No. of injuries | % |
| --- | --- | --- |
| **Digits** | 412 | 33.8 |
| Shoulder | 374 | 30.7 |
| Elbow | 213 | 17.5 |
| Wrist | 176 | 14.4 |
| Palm | 44 | 3.6 |

Values are presented as n (%).
All injuries were acute sports-related upper-extremity injuries. Only one injury per athlete was included in the analysis. Digits injuries included the thumb and fingers distal to the metacarpophalangeal joint.

### Anatomical Characteristics of Digit Injuries

Among the 412 digit injuries, analysis at the digit level showed that the thumb (109 cases, 26.4%) and little finger (97 cases, 23.5%) were most frequently affected (Table 2A).

**Table 2.** Anatomical distribution of digits injuries (n = 412)

|  |  |
| --- | --- |
| A. Digit-level distribution |  |
| Thumb | 109 (26.4%) |
| Index | 61 (14.8%) |
| Middle | 69 (16.7%) |
| Ring | 76 (18.4%) |
| Little | 97 (23.5%) |

B. Joint-level distribution by digit group
| Thumb (n=109) |  |
| --- | --- |
| IP | 24 (22.0%) |
| MP | 79 (72.5%) |
| Others | 6 (5.5%) |

| Fingers (n=303) |  |
| --- | --- |
| DIP | 40 (13.2%) |
| PIP | 182 (60.1%) |
| MP | 55 (18.2%) |
| Others | 26 (8.6%) |

Digits injuries were defined as injuries involving the digits distal to the metacarpophalangeal (MP) joint, including the MP joint itself. Injuries involving the palm or wrist were excluded and classified separately.

At the joint level, MP joint injuries accounted for 72.5% of thumb injuries, whereas PIP joint injuries were most common in the other digits, accounting for 60.1% of cases (Table 2B).

### Injury Types, Mechanisms, and Sports Categories

Regarding injury type, jammed finger was the most frequent diagnosis, accounting for 267 cases (64.8%), followed by fractures (83 cases, 20.1%) and dislocations (22 cases, 5.3%).

Most injuries were caused by contact mechanisms (90.3%), with ball contact (49.5%) and player-to-player contact (28.2%) being the most common specific causes. Ball sports accounted for the majority of injuries (85.4%) (Table 3).

**Table 3.** Injury characteristics and mechanisms of digits injuries (n = 412)

| A. Injury type |  |
| --- | --- |
| Fracture | 83 (20.1%) |
| Dislocation | 22 (5.3%) |
| Jammed finger | 267 (64.8%) |
| Others | 40 (9.7%) |

| B. Injury Mechanism |  |
| --- | --- |
| Contact | 372 (90.3%) |
| Non-contact | 40 (9.7%) |

| **C. Specific cause** |  |
| --- | --- |
| Ball contact | 204 (49.5%) |
| Player contact | 116 (28.2%) |
| Others | 92 (22.3%) |

| **D. Sports category** |  |
| --- | --- |
| Ball sports | 352 (85.4%) |
| Combat sports | 38 (9.2%) |
| Others | 22 (5.3%) |

Jammed finger includes ligamentous injury without radiographic fracture.
Volar plate injuries with radiographic avulsion fragments were classified as fractures.

## Discussion

### Principal Findings

In this study, we analyzed 1,219 acute sports-related upper-extremity injuries in athletes aged ≤22 years and demonstrated that digit injuries were the most common injury location, accounting for 412 cases (33.8%). This frequency exceeded that of major joints such as the shoulder, elbow, and wrist, highlighting the substantial clinical burden of digit injuries in young athletes (1,2).

Furthermore, jammed finger was the most frequent injury type, accounting for 64.8% of digit injuries, occurring more often than fractures or dislocations. Most injuries were caused by contact mechanisms, with ball contact being the predominant cause. These findings suggest that digit injuries in young athletes are closely associated with sport-specific injury environments (4,5).

### Comparison With Previous Studies

Previous epidemiological studies on digit injuries have primarily focused on general trauma populations or patients presenting to emergency departments, and reports specifically addressing sports-related digit injuries are limited (3,4). In these studies, digit injuries have been reported to account for approximately 15–30% of all injuries; however, non–sports-related injuries and adult populations were often included (3,4).

A key feature of the present study is that the denominator was restricted to acute sports-related upper-extremity injuries in athletes aged ≤22 years. Under this condition, digit injuries emerged as the most frequent injury type. This finding likely reflects the vulnerability of the digits to direct external forces during sports activities (1,2).

In addition, whereas fractures have frequently been reported as a major component of digit injuries in previous studies (3,4), jammed finger constituted the majority of injuries in the present cohort. This difference may be attributable to differences in study populations as well as the high prevalence of mild to moderate ligamentous and capsular injuries characteristic of sports-related trauma (5,6).

### Anatomical and Functional Characteristics of Digit Injuries

At the digit level, injuries to the thumb and little finger were most common. The thumb plays a critical role in grip and ball control, whereas the little finger is located on the ulnar side of the hand and is more susceptible to contact and collision during sports participation. These anatomical and functional characteristics may explain the observed digit-specific injury distribution (5).

At the joint level, MP joint injuries were most common in the thumb, whereas PIP joint injuries predominated in the other digits. The PIP joint is structurally vulnerable to valgus and hyperextension stresses and is known to be susceptible to injury during ball contact or collision (5,6). The present findings are consistent with these biomechanical characteristics.

### Clinical Implications and Preventive Considerations

The present results indicate that digit injuries, particularly jammed finger, occur at a frequency that should not be overlooked in young athletes (4,5). Although jammed finger is often regarded as a relatively minor injury, inappropriate initial management or continued sports participation may result in chronic joint instability or persistent range-of-motion limitations (6).

Currently, standardized guidelines for the treatment and rehabilitation of jammed finger are lacking, and management strategies often rely on immobilization-based approaches (5,6). Such strategies may contribute to joint stiffness and delayed functional recovery, representing an ongoing clinical challenge.

Given that most injuries were caused by ball contact, preventive strategies such as sport-specific finger protection, taping, and technical instruction may be effective in reducing injury risk (1,2,5). In addition, management strategies emphasizing appropriate early assessment and timely initiation of range-of-motion exercises may facilitate functional recovery. Further clinical studies are warranted to validate these approaches.

### Limitations

This study has several limitations. First, as a single-center retrospective study, the findings may be influenced by regional factors and the distribution of sports activities (1,2). Second, injury severity, treatment outcomes, and time to return to sport were not evaluated. Third, the diagnosis of jammed finger was based on clinical assessment and plain radiographs to exclude fractures; however, detailed ligament-specific subclassification using advanced imaging modalities such as MRI was not performed (5,6).

## Conclusion

In athletes aged ≤22 years, digit injuries were the most frequent type of acute sports-related upper-extremity injury, with jammed finger accounting for the majority of cases. Most injuries were caused by ball contact, underscoring the importance of preventive strategies and appropriate initial management for digit injuries in young athletes.

These findings emphasize that digit injuries, often regarded as minor, represent a substantial clinical burden in youth sports and warrant greater attention in both prevention and early management.

## Data Availability

All data produced in the present study are available upon reasonable request to the authors

## Declaration of generative AI and AI-assisted technologies in the manuscript preparation process

During the preparation of this work, the author used ChatGPT (OpenAI) for language editing and improvement of clarity. After using this tool, the author reviewed and edited the content as needed and takes full responsibility for the content of the published article.

